# Modelling the COVID-19 Fatality Rate in England and its Regions

**DOI:** 10.1101/2021.01.19.21249816

**Authors:** Nigel Saunders

## Abstract

A model to account for the fatality rate in England and its regions is proposed. It follows the clear observation that, rather than two connected waves, there have been many waves of infections and fatalities in the regions of England of various magnitudes, usually overlapping. The waves are self-limiting, in that clear peaks are seen, particularly in reported positive test rates. The present model considers fatalities as the data reported are more reliable than positive test rates, particularly so during the first wave when so little testing was done.

The model considers the observed waves are essentially similar in form and can be modelled using a single wave form, whose final state is only dependent on its peak height and start date. The basic wave form was modelled using the observed fatality rates for London, which unlike the other regions, exhibited almost completely as a single wave in the “first wave”. Its form matches rather well with the “Do Nothing” model reported by Imperial College on 16^th^ March 2020, but reduced substantially from its expected peak.

There are, essentially, only two adjustable parameters used in the model, the start date of the relevant wave and its height. The modelled fatalities for each wave are summated per day and a cumulative curve is matched to that reported. The minimal number of adjustable parameters, alongside the fact that the waves invariably overlap, provides highly stringent conditions on the fitting process.

Results are presented for each region for both the “first” and “second’ waves. High levels of accuracy are obtained with R^2^ values approaching 100% against the ideal fit for both waves. It can also be seen that there are fundamental differences between the underlying behaviour of the “first” and “second” waves and reasons as to why those differences have arisen is briefly discussed.

## 1. Introduction

While there are daily reports of the progress of COVID-19 infections in the UK, it seems little work has been done in trying to understand how the virus has actually spread and grown in the UK. Regional R rates are regularly reported, however, infection or fatality rates over the whole period for the regions are rarely, if ever presented. By doing so, a far more detailed view of how the infection has spread and grown is revealed. Fig.1 below shows population adjusted daily positive tests for the regions of England from the beginning of the pandemic using data reported up to 1^st^ March 2021 ^(1)^. A 7-day average of cases by specimen date (*averaged at midweek*) is used and the last 5 days not used due to likely incompleteness as advised by the GOV.uk site.

**Fig. 1.**
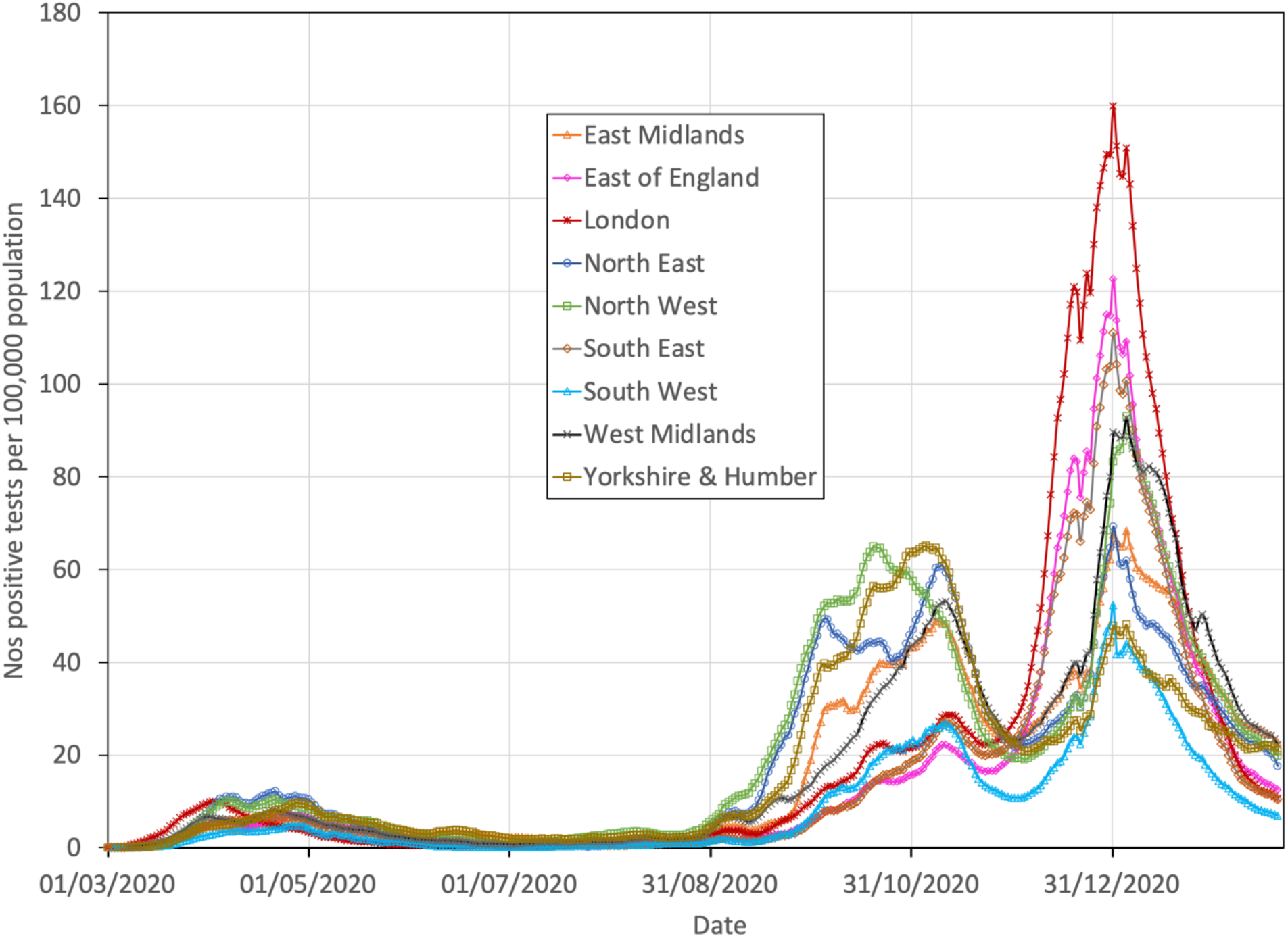
Reported population adjusted daily positive tests for the regions of England from data reported from the beginning of the pandemic, up to 1^st^ March 2021 ^(1)^ using a 7-day average of cases by specimen date

Examination of Fig.1 provides clear light on how the virus has spread and grown across England. (Note, the variability in the curves at the end of December and beginning of January can be explained by under recording around Christmas and New Year’s day.)

- There have been many waves of infection in the regions of various magnitudes, many major, but with some small but distinctly observable.
- In the “first wave”, with the exception of London, which exhibited as an essentially singular wave, multiple overlapping waves can be seen in the other regions, exhibiting as multiple peaks within a broad “first” regional wave.
- After the first lockdown, infections continued to fall in a quite continuous fashion. There were some minor waves but, effectively, rates plateaued, showing only a slight rise until week 35, after which rates increased in all regions nationwide.
- Since week 35 there appears to be multiple overlapping, but distinct waves in all of the regions. While some waves are clearly apparent, they can also be seen as inflections in the slope of the test data, which occur as an increasingly dominant wave adds to the infection curve.
- During the second week of October, with the exception of the North West which peaked in the second half of September, there was a fall in infection rates for all regions, with the Northern and Midland regions exhibiting particularly sharp falls. Subsequently, around the end of November/beginning of December, infection rates rose in all regions, but most sharply in London, the South East and the East of England. All regions subsequently peaked at the end of December/beginning of January before rates fell sharply.

### 2. Development of the model

From Fig.1, London is the only region that exhibited as mainly a single wave during the “first wave”, which is rather symmetrical in form, and can be used a basis for developing a model consisting of multiple waves. Rather than using positive test data, the model has been developed taking as its basis fatality rates. The data is far more reliably reported over the whole period and the reported test data also does not accurately reflect the true total number of infections due to asymptomatic cases.

Fig.2 shows reported fatalities by date of death ^(1)^, as a 7-day average for London. The wave is quite symmetrical in nature until the 29^th^ April, at which point a further wave becomes apparent, adding to the first, before diminishing to almost zero. The model used here fits the curve up until then, before itself diminishing to zero (Appendix 1).

**Fig. 2.**
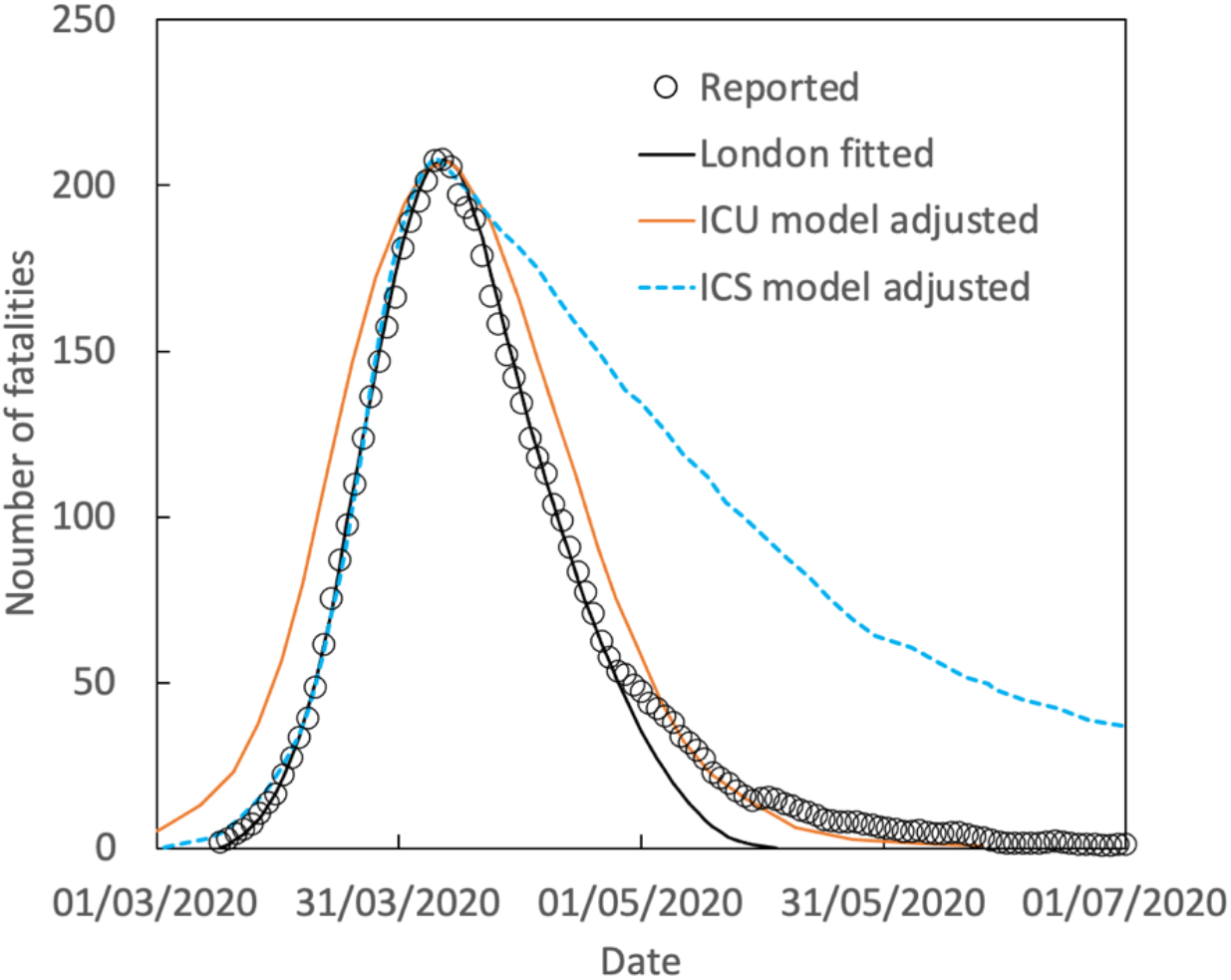
Fatality rates reported for London including those from the present model and using rate adjusted ICU and ICS models.

Fig.2 also includes two further curves taken after the Imperial College report of 16^th^ March 2020 ^(2)^.

1. A curve based on the “Do Nothing” scenario (hereafter called the Imperial College Unconstrained (ICU) model), scaled to match the peak height of the reported curve for London and date adjusted.
2. A curve based on the scenario of Schools and Universities Closure, Case Isolation and Population-Wide Social Distancing (hereafter called the Imperial College Suppression (ICS) model). The reported curve is provided in terms of critical care beds occupied and, based on the assumption it is directly proportional to subsequent fatalities, scaled to match the peak height for London and date adjusted.

The scaled ICU curve matches the reported data for the first wave rather well, though is a little wider, while the ICS curve matches the reported data almost exactly to the peak height before markedly diverging.

The fitted curve can now be used in a multiple wave model which requires three sets of input. The first is the number of waves, which is usually clear to see or becomes required during the fitting process. Otherwise, the fitting process requires only two parameters (i) the start date of the relevant wave and (ii) its height. The calculated fatalities for each wave are then summated and the cumulative curve matched to the reported data. The model was implemented here using spreadsheets but should suit a computer programme utilising a least squares methodology to minimise the error.

## 3 Results

### 3.1 “First Wave”

Figs. 3-11(a&b) show results for the fitted curves to reported data alongside plots for the divergence from the “ideal fit” in terms of the value of R^2^. The data is given as fatalities, reported by date of death ^(1)^, as a 7-day average.

**Fig 3.**
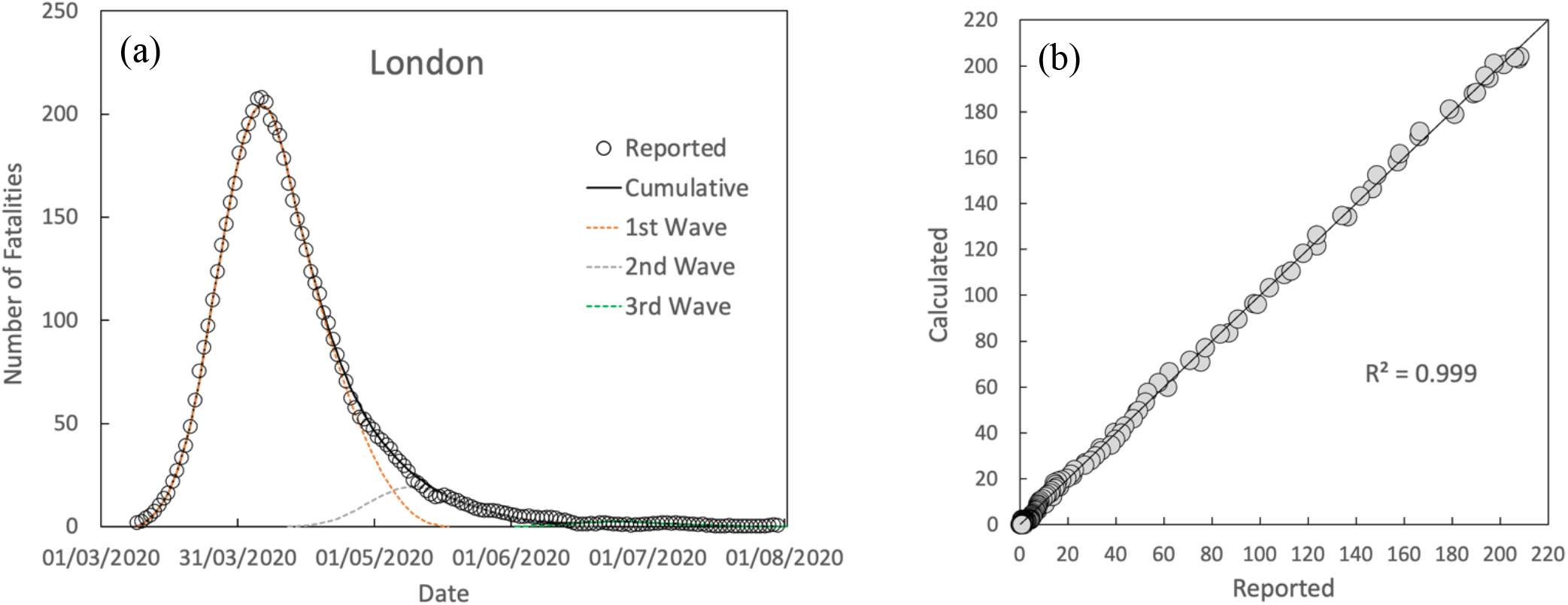
(a) Comparison between calculated and reported fatalities by date of death (7-day average) and (b) divergence from the “ideal fit” for London in the “first wave”

**Fig. 4.**
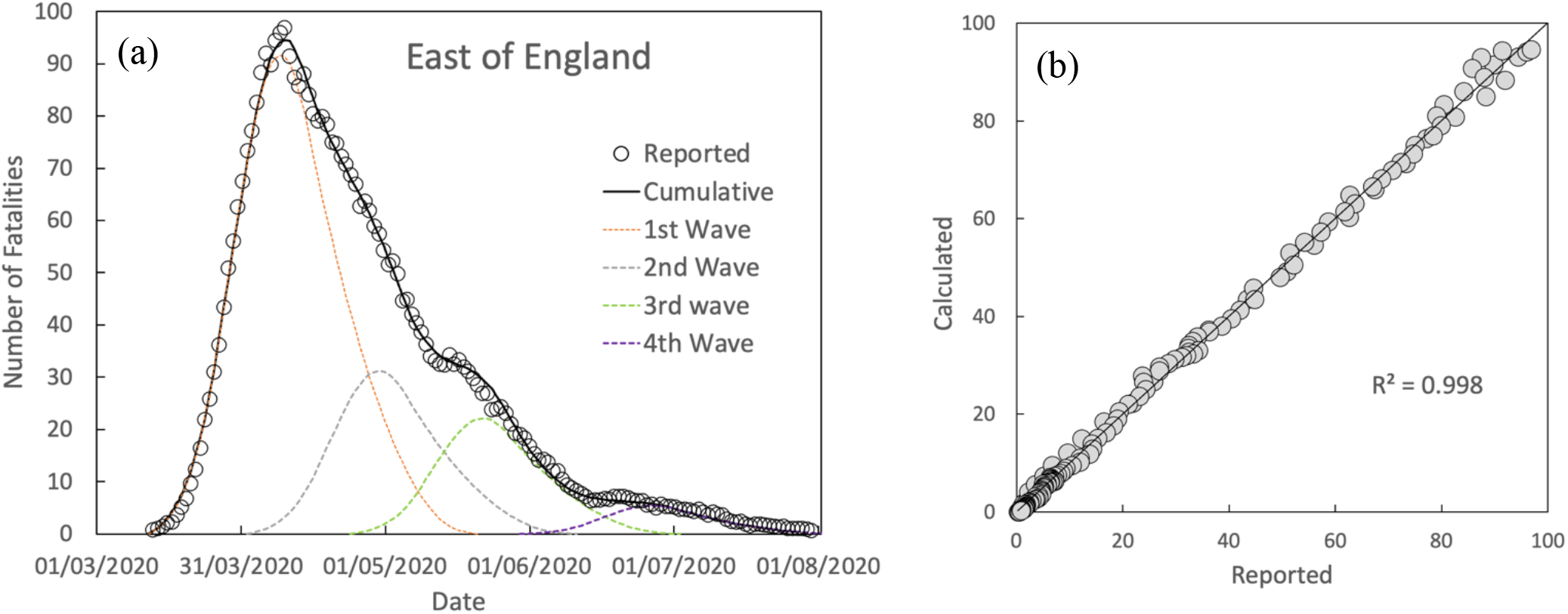
(a) Comparison between calculated and reported fatalities by date of death (7-day average) and (b) divergence from the “ideal fit” for the East of England in the “first wave”

**Fig. 5.**
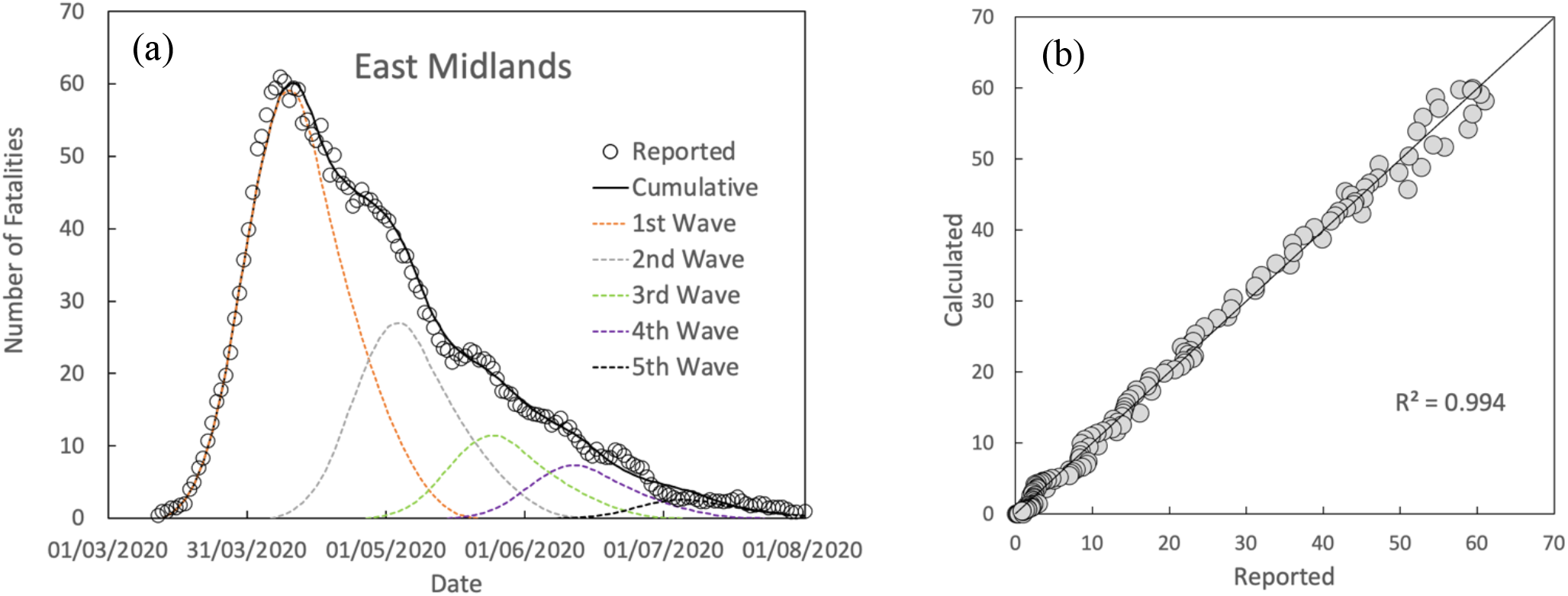
(a) Comparison between calculated and reported fatalities by date of death (7-day average) and (b) divergence from the “ideal fit” for the East Midlands in the “first wave

**Fig. 6.**
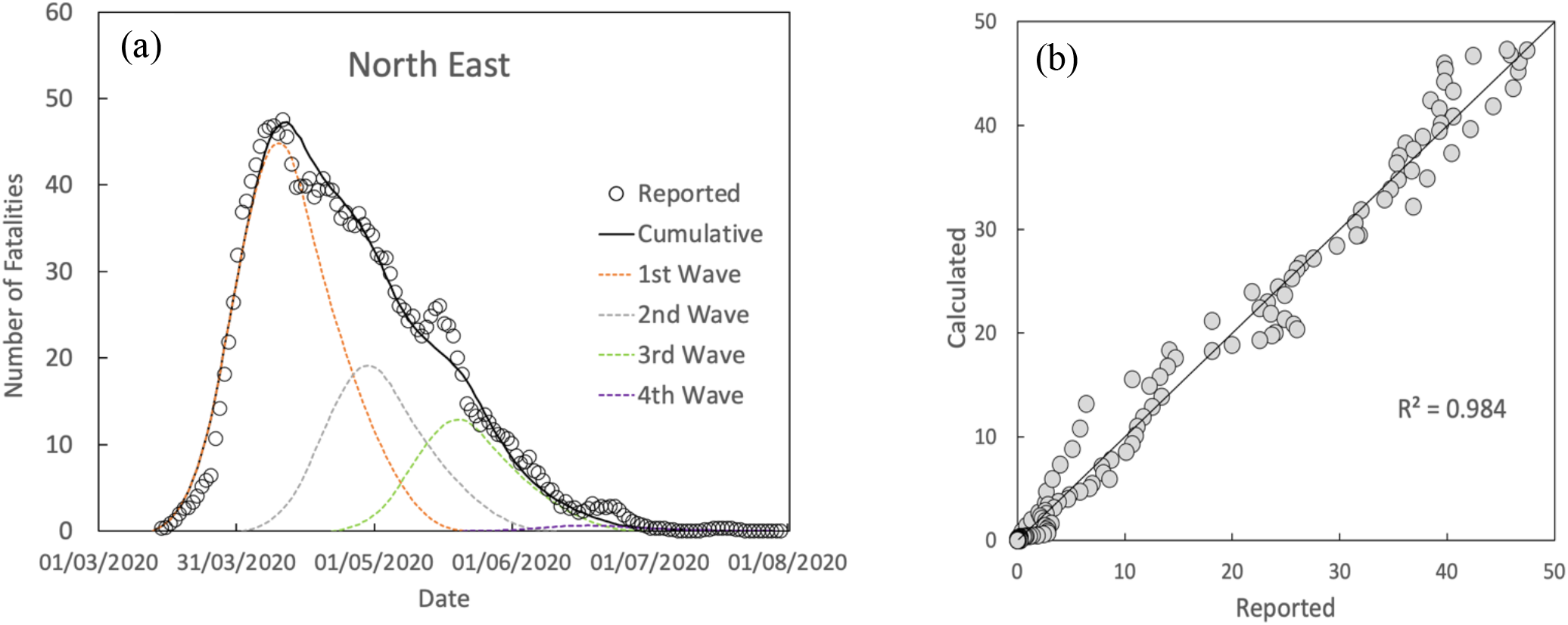
(a) Comparison between calculated and reported fatalities by date of death (7-day average) and (b) divergence from the “ideal fit” for the North East in the “first wave”

**Fig. 7.**
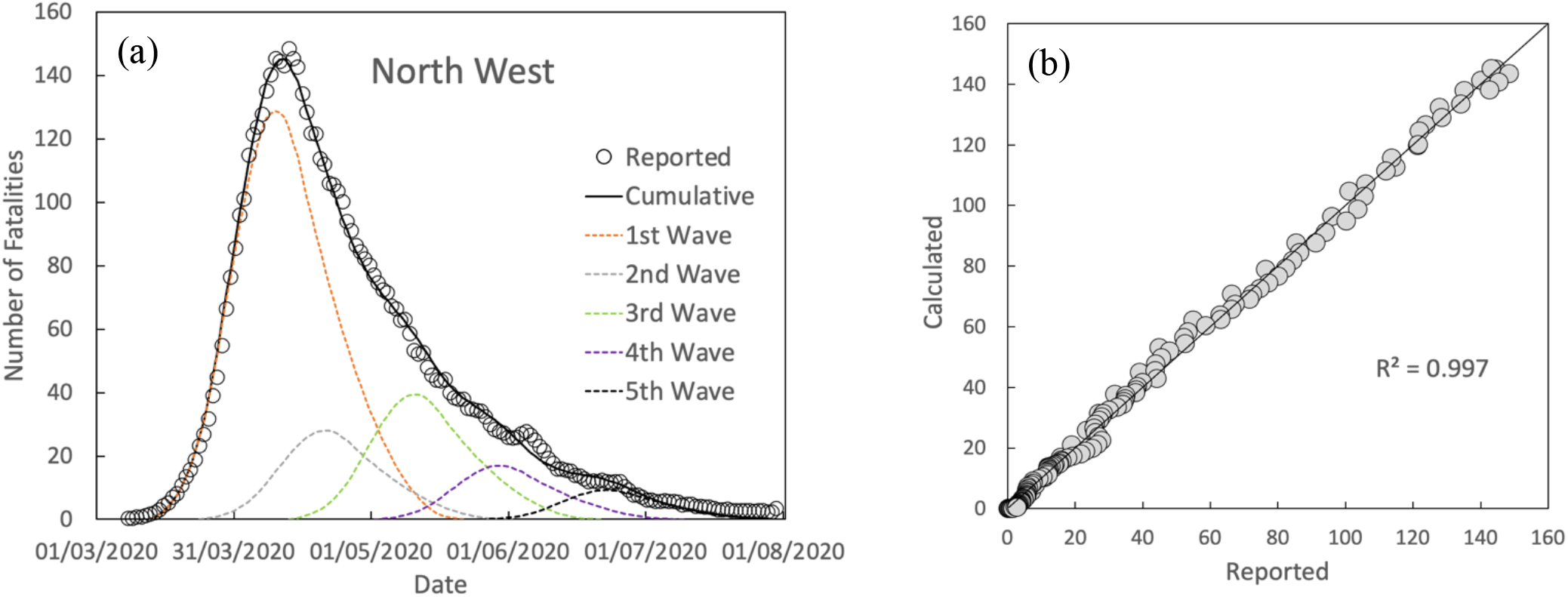
(a) Comparison between calculated and reported fatalities by date of death (7-day average) and (b) divergence from the “ideal fit” for the North West in the “first wave”

**Fig. 8.**
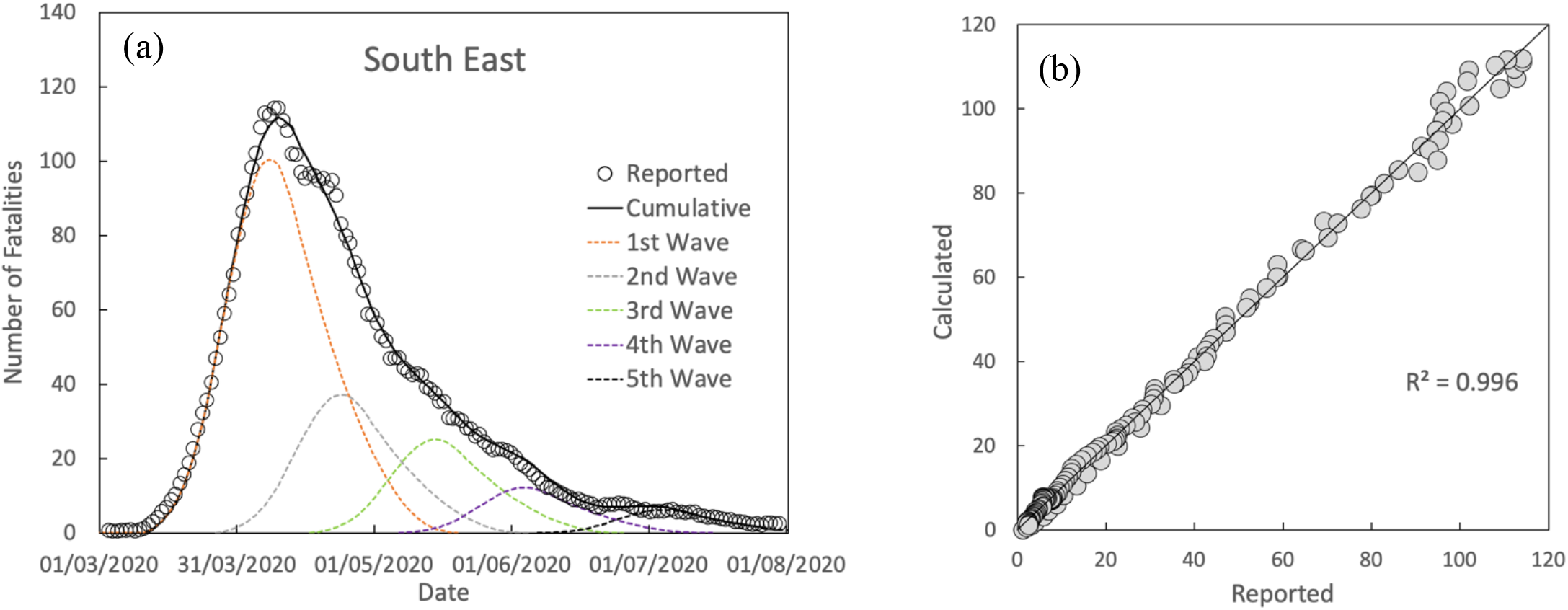
(a) Comparison between calculated and reported fatalities by date of death (7-day average) and (b) divergence from the “ideal fit” for the South East in the “first wave”

**Fig. 9.**
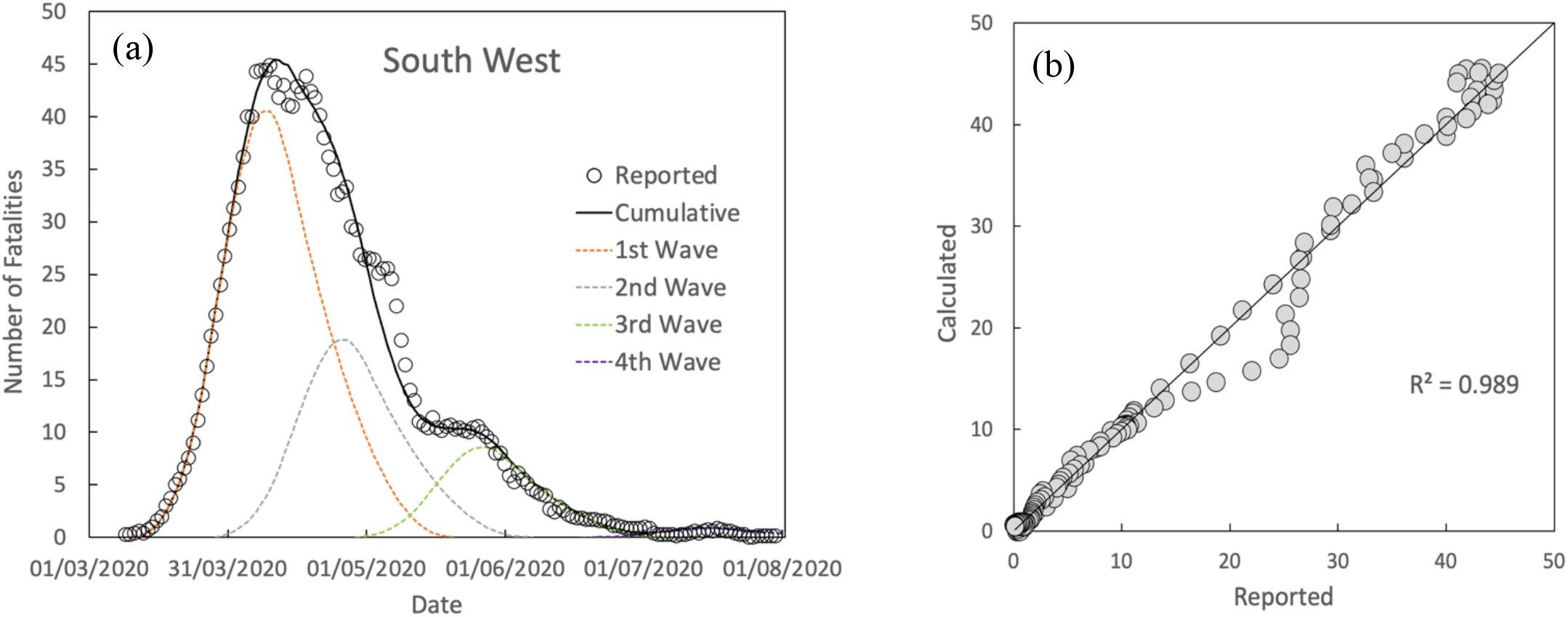
(a) Comparison between calculated and reported fatalities by date of death (7-day average) and (b) divergence from the “ideal fit” for the South West in the “first wave”

**Fig. 10.**
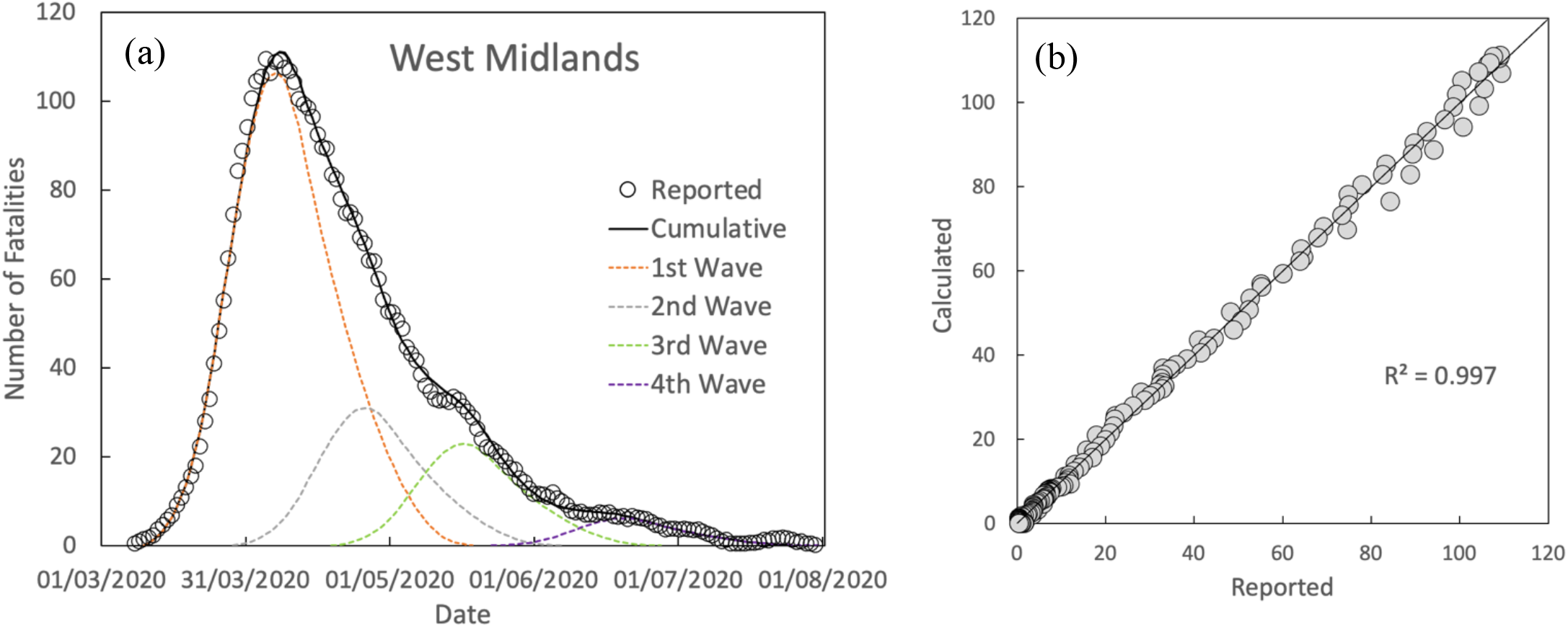
(a) Comparison between calculated and reported fatalities by date of death (7-day average) and (b) divergence from the “ideal fit” for the West Midlands in the “first wave”

**Fig. 11.**
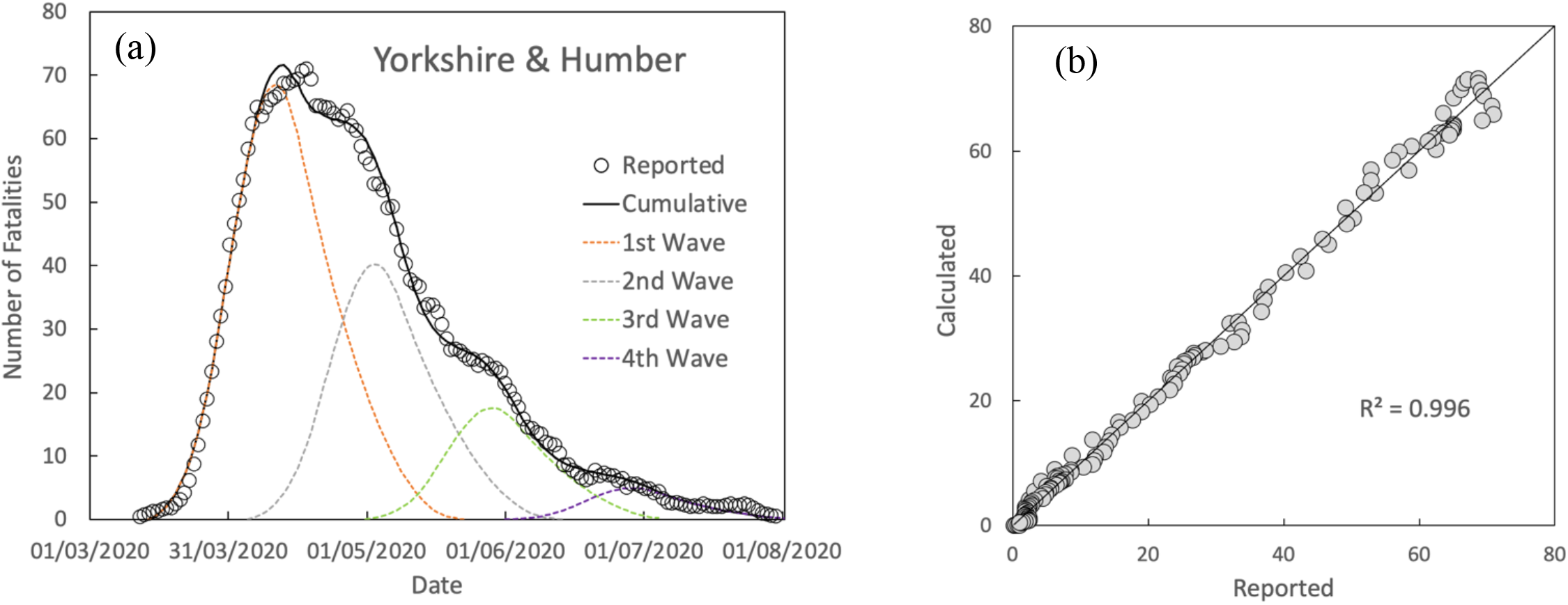
(a) Comparison between calculated and reported fatalities by date of death (7-day average) and (b) divergence from the “ideal fit” for Yorkshire & Humber in the “first wave”

Considering the simplicity of the model, it matches the general behaviour of fatality rates very accurately, but there are some spikes in fatalities which cannot be matched. It may be, straightforwardly, that the model is unable to account for minor variations in behaviour or due to clusters of fatalities in settings where fatality rates would be higher than for the overall population average.

Fig. 12 shows the sum of all the waves of the regions compared to fatality data reported for England ^(1)^ as a 7-day average, while Fig.13 compares the raw data as reported by date of death for completeness.

**Fig 12.**
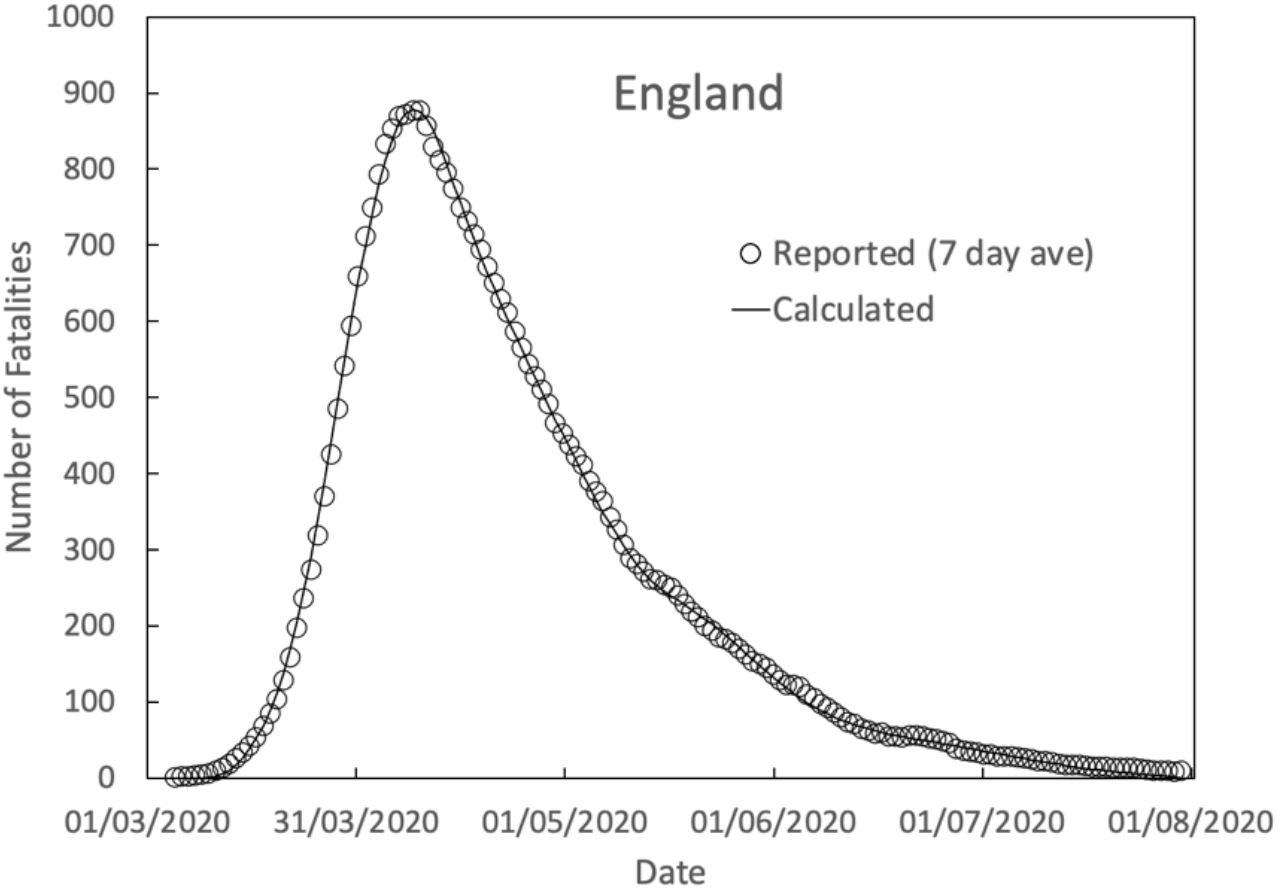
Comparison between the sum of all calculated waves of the regions compared to reported fatalities by date of death (7-day average) for England during the “first wave”

**Fig. 13.**
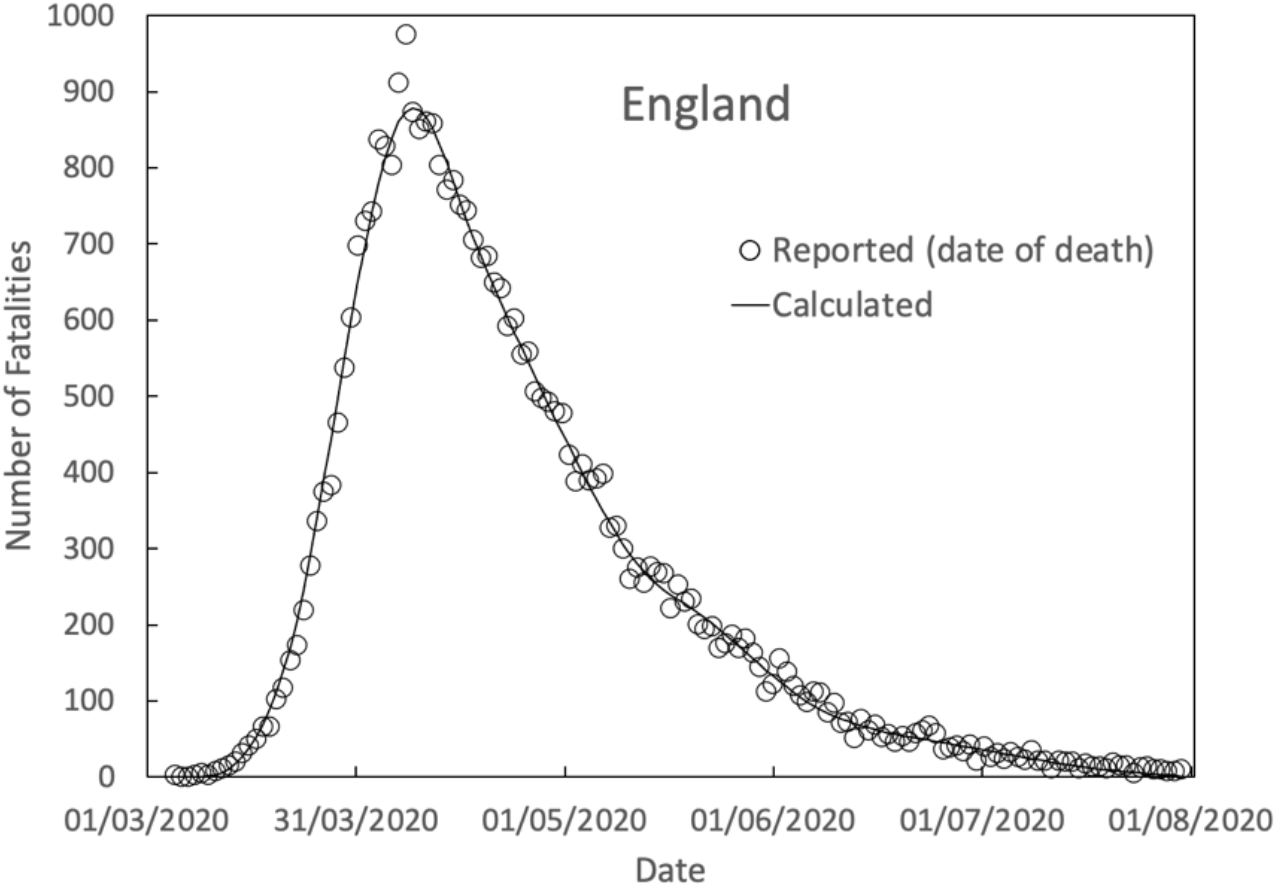
Comparison between the sum of all calculated waves of the regions compared to reported fatalities by date of death for England during the “first wave”

### 3.2 “Second Wave”

Figs. 14-22 (a&b) show, similarly to the “first wave”, results for the fitted curves to data reported up until 1^st^ March ^(1)^ alongside plots for the divergence from the “ideal fit” in terms of the value of R^2^. As for Fig.1 the last 5 days of the reported data are not considered. However, it is noted that there is significant delay in reporting, with fatality results still being reported after the 5-day delay accounted for here. The reported and subsequently fitted rates for the later dates, therefore, have some uncertainty. However, there appears to be a clear trend in London, the East of England and the South East for the most recently reported fatalities to fall below the calculated line which could be taken as a clear sign that the major vaccination drive in the UK is having an effect and suppressing fatalities during the latest wave in these regions.

**Fig 14.**
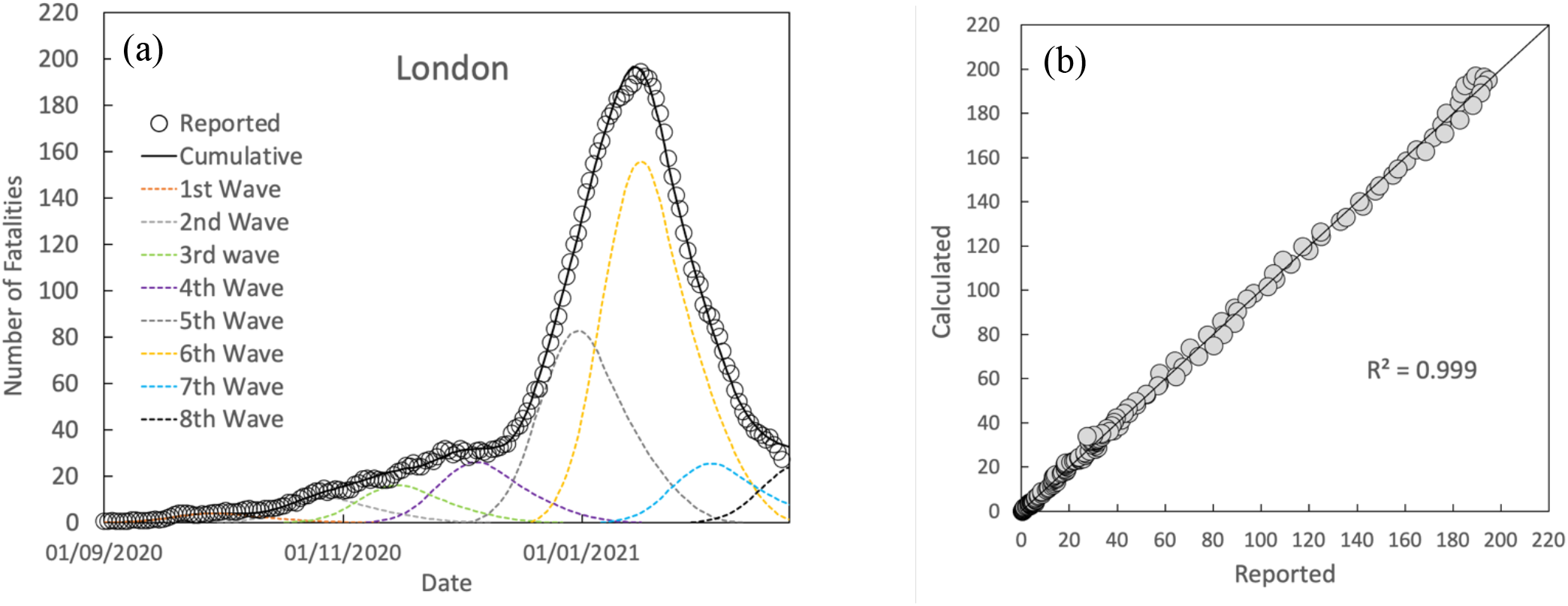
(a) Comparison between calculated and reported fatalities by date of death (7-day average) and (b) divergence from the “ideal fit” for London in the “second wave”

**Fig. 15.**
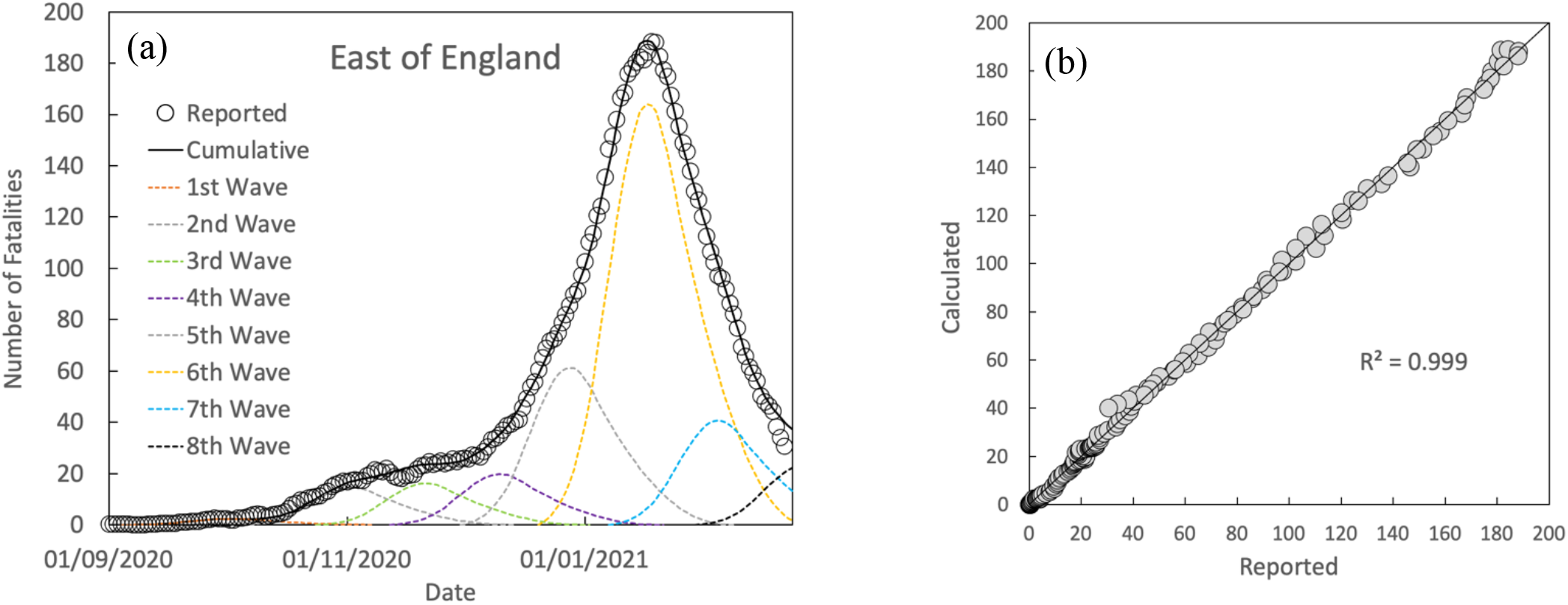
(a) Comparison between calculated and reported fatalities by date of death (7-day average) and (b) divergence from the “ideal fit” for the East of England in the “second wave”

**Fig. 16.**
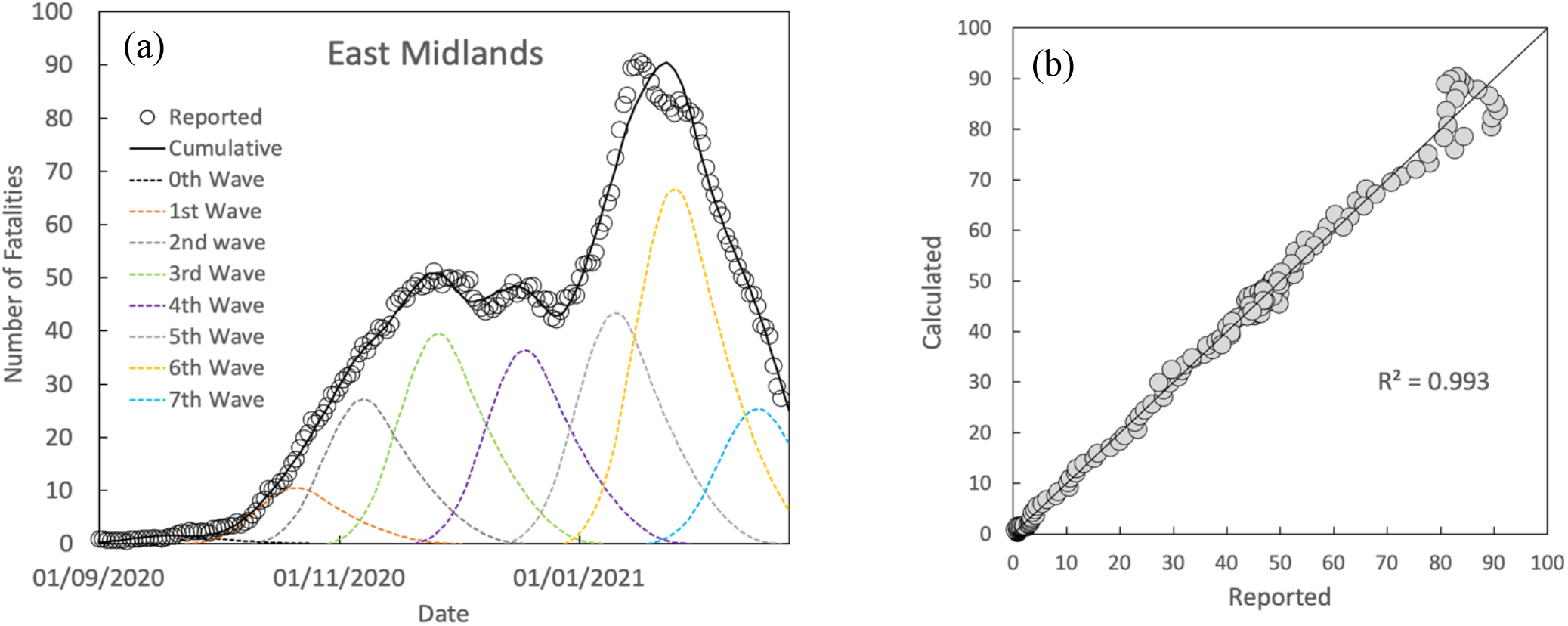
(a) Comparison between calculated and reported fatalities by date of death (7-day average) and (b) divergence from the “ideal fit” for the East Midlands in the “second wave”

**Fig. 17.**
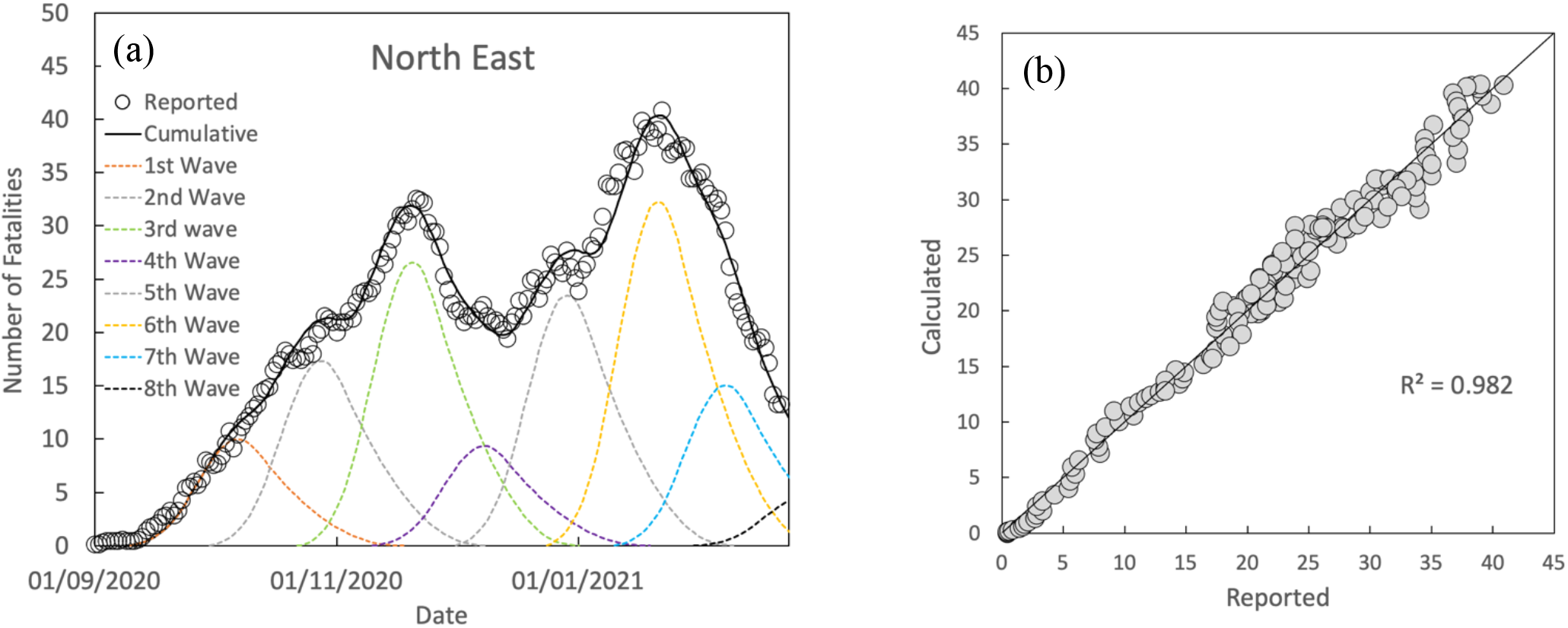
(a) Comparison between calculated and reported fatalities by date of death (7-day average) and (b) divergence from the “ideal fit” for the North East in the “second wave”

**Fig. 18.**
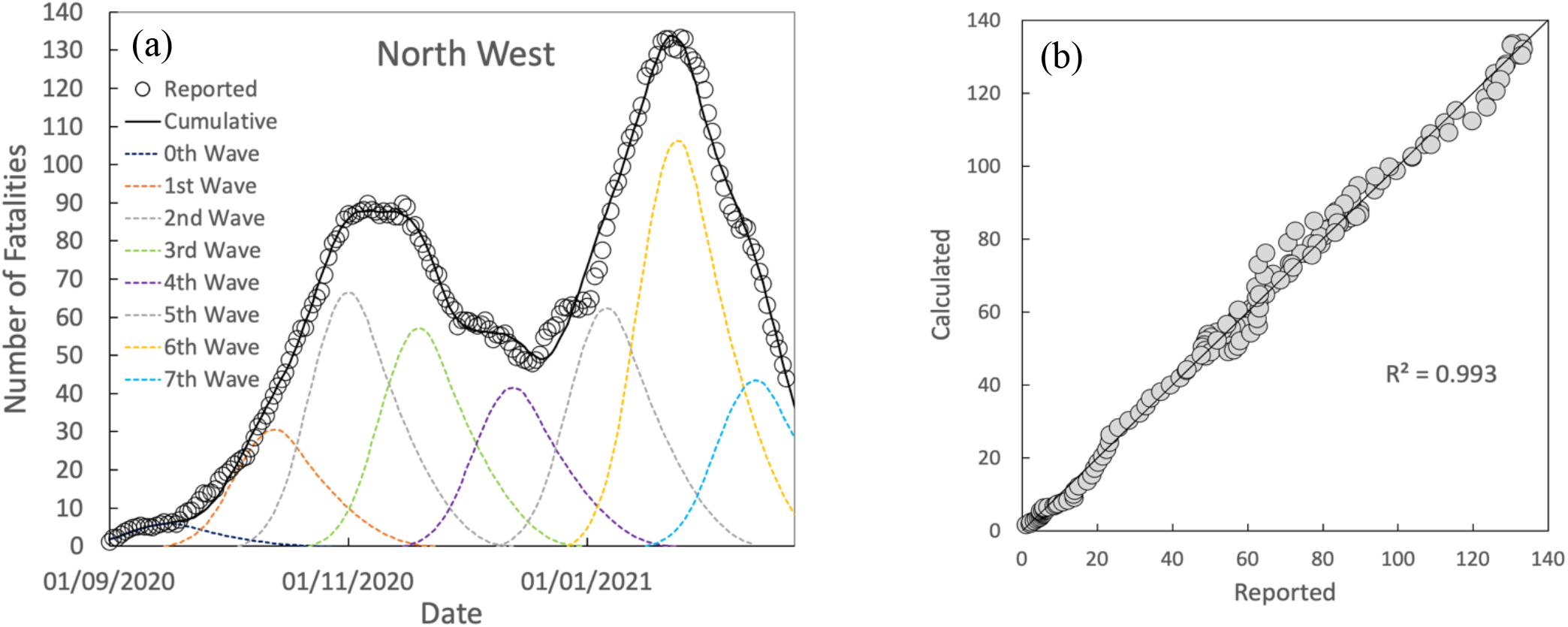
(a) Comparison between calculated and reported fatalities by date of death (7-day average) and (b) divergence from the “ideal fit” for the North West in the “second wave”

**Fig. 19.**
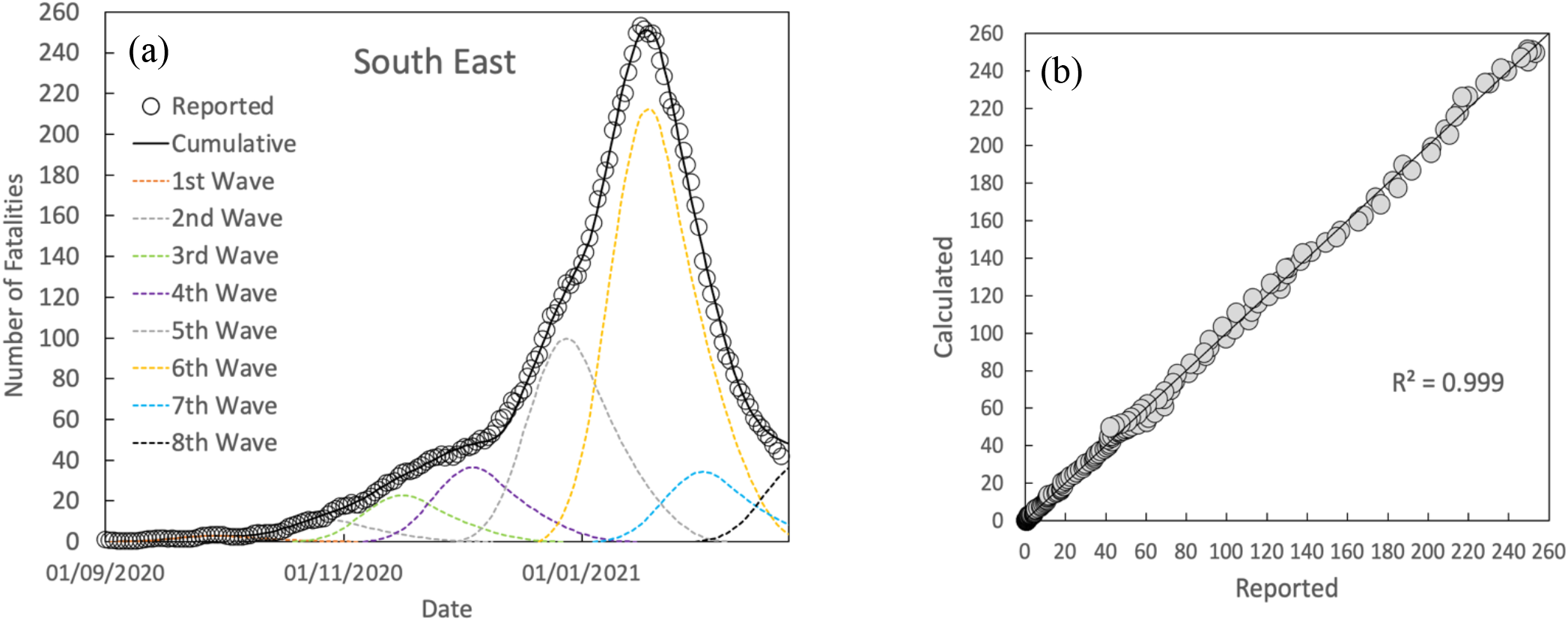
(a) Comparison between calculated and reported fatalities by date of death (7-day average) and (b) divergence from the “ideal fit” for the South East in the “second wave”

**Fig. 20.**
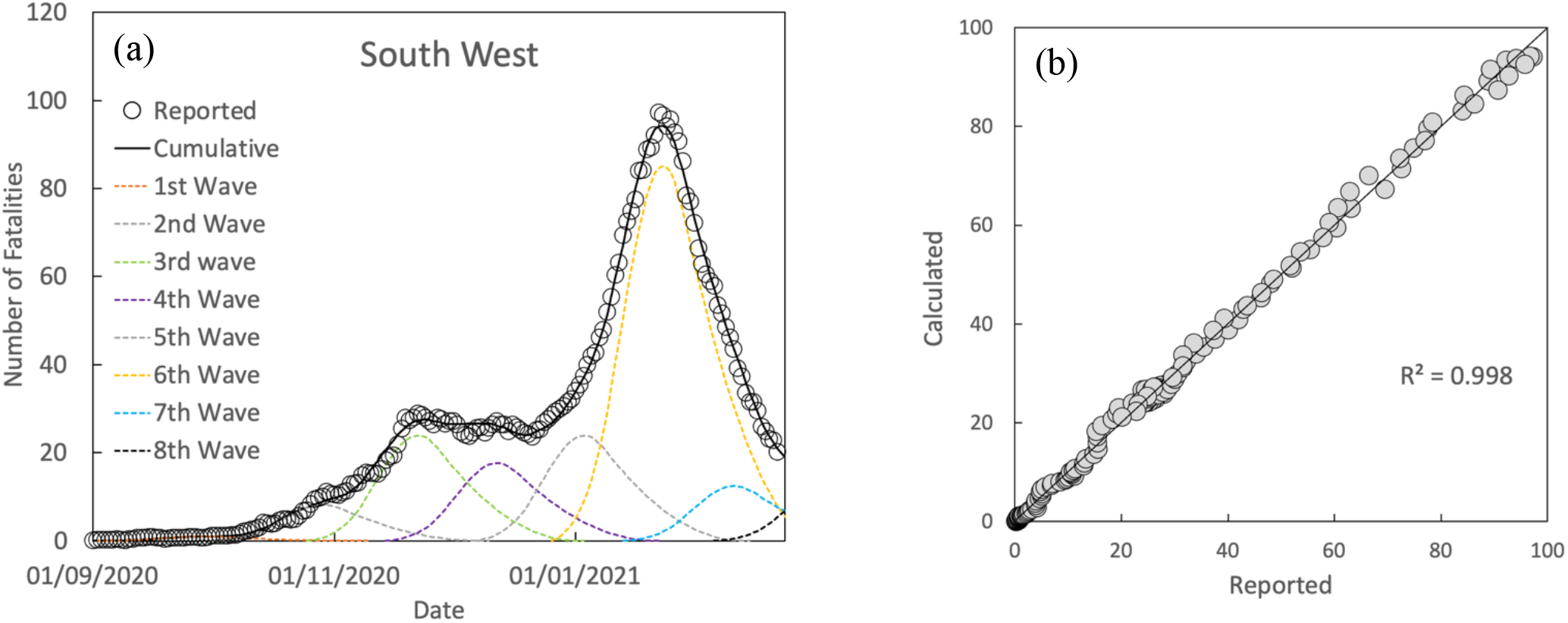
(a) Comparison between calculated and reported fatalities by date of death (7-day average) and (b) divergence from the “ideal fit” for the South West in the “second wave”

**Fig. 21.**
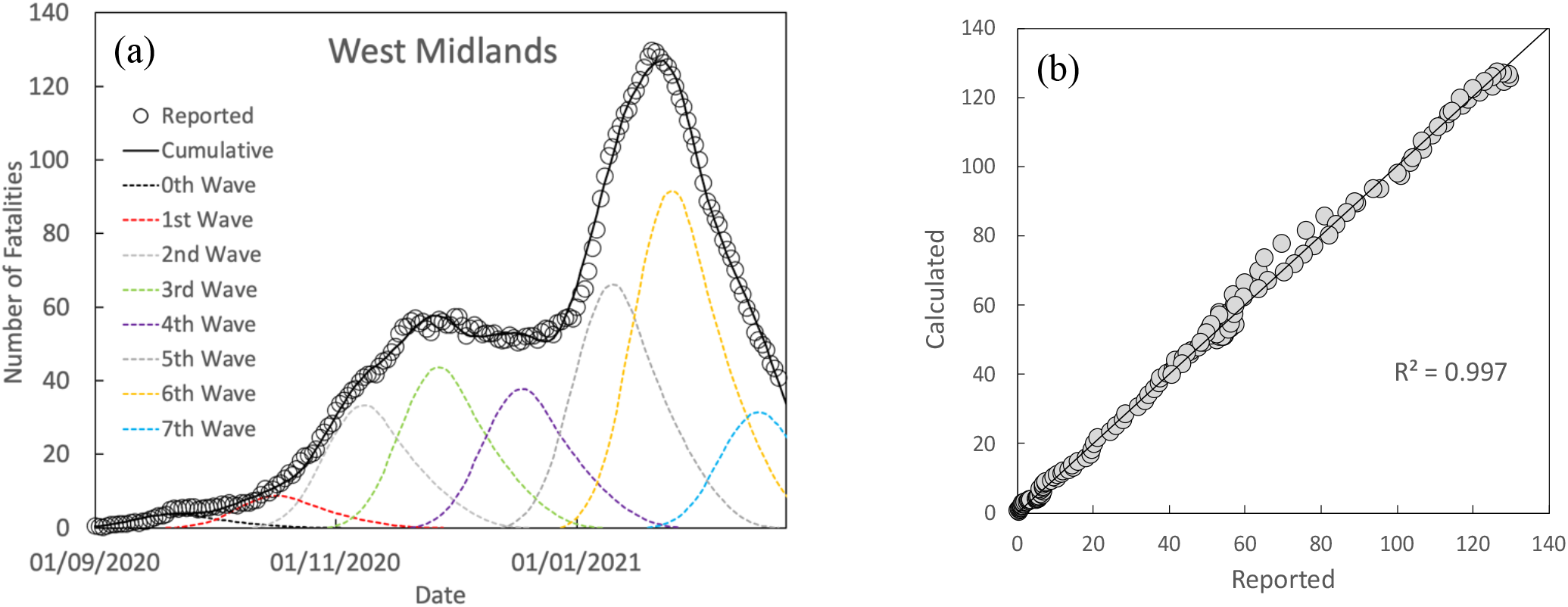
(a) Comparison between calculated and reported fatalities by date of death (7-day average) and (b) divergence from the “ideal fit” for the West Midlands in the “second wave”

**Fig. 22.**
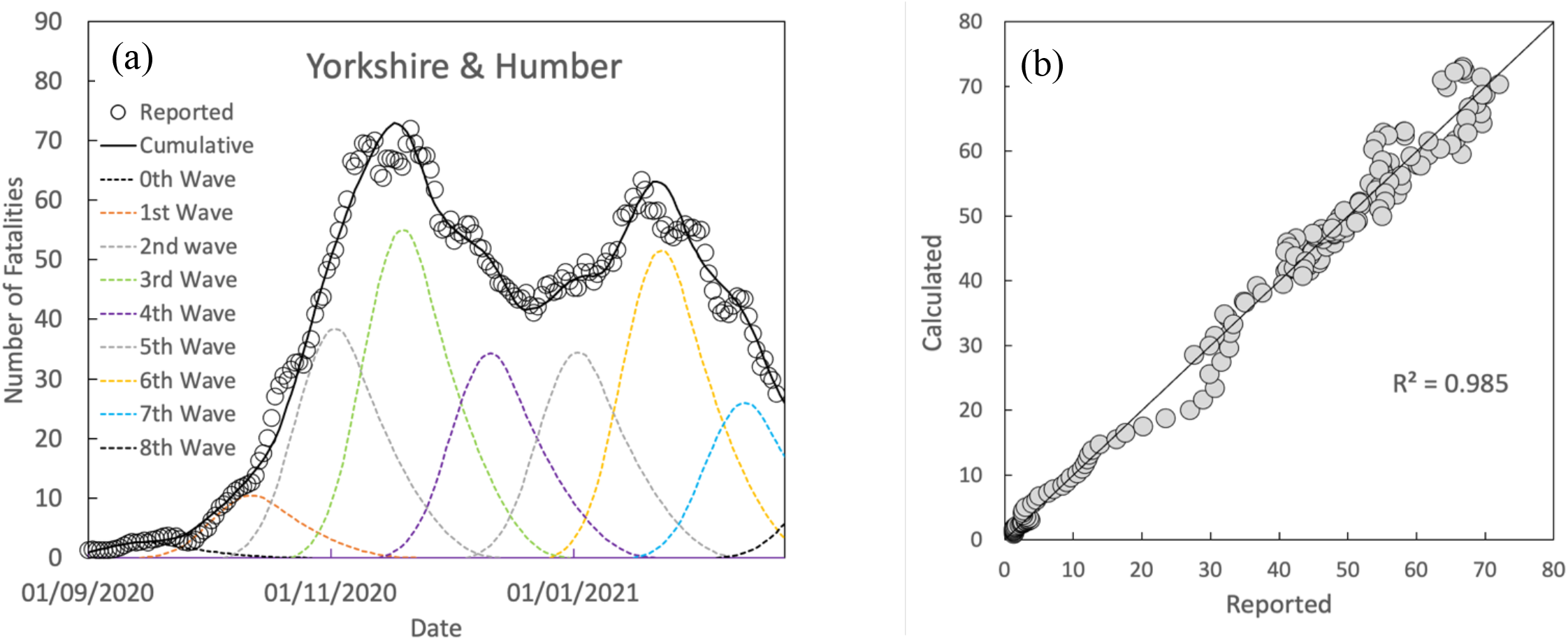
(a) Comparison between calculated and reported fatalities by date of death (7-day average) and (b) divergence from the “ideal fit” for the West Midlands in the “second wave”

The numbering of the waves follows the convention whereby, the 1^st^ Wave corresponds to the first wave that started during September, while the 0^th^ Wave corresponds to a wave that began in August. Fig.23 show the sum of all the waves of the regions compared to fatality data reported for England ^(1)^ as a 7-day average, while Fig.24 compares the raw data as reported per day for completeness. All regions show at least 7 waves from the beginning of September with a further 8^th^ wave appearing in some. A further common feature for all regions is the periodic nature of the waves.

**Fig. 23.**
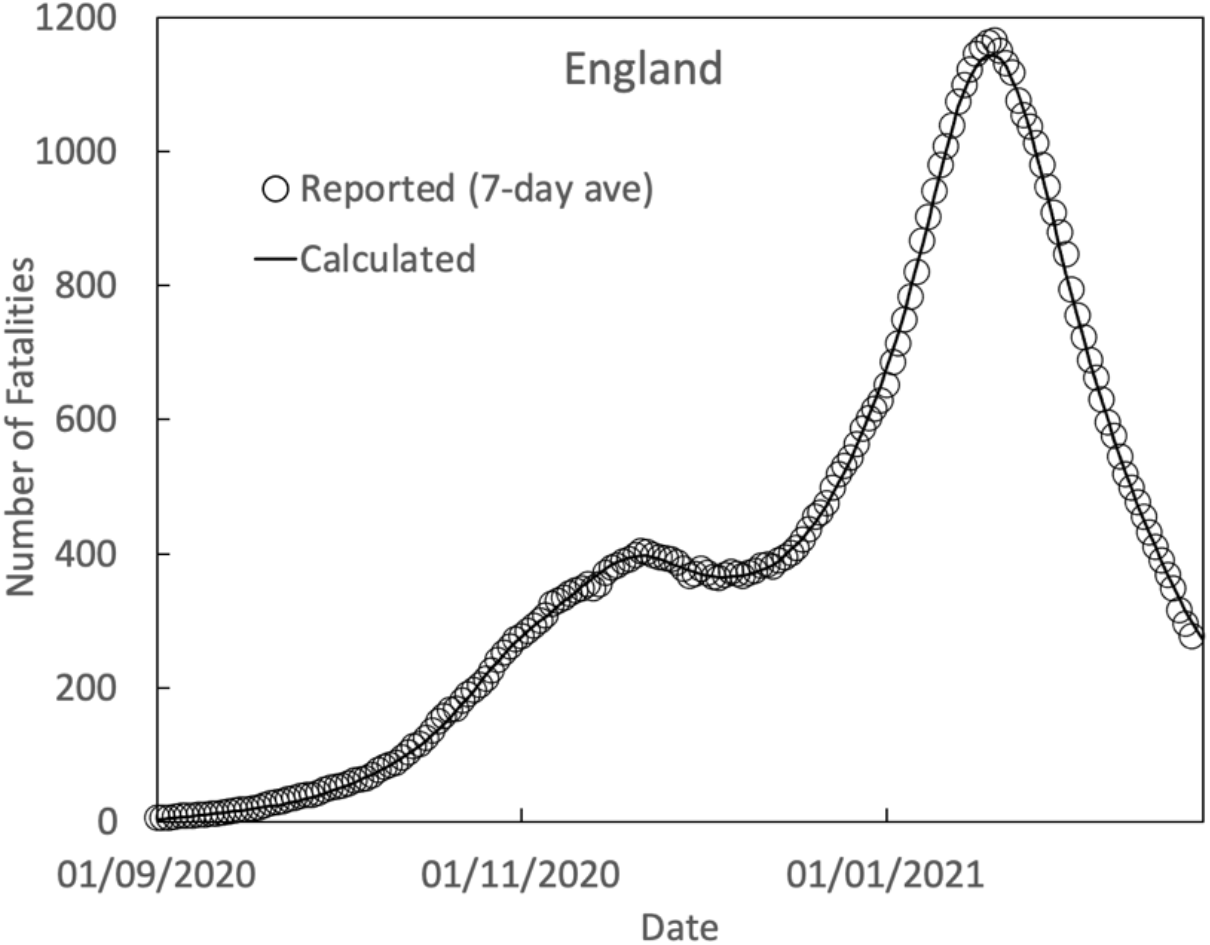
Comparison between the sum of all calculated waves of the regions compared to reported fatalities by date of death (7-day average) for England during the “second wave”

**Fig. 24.**
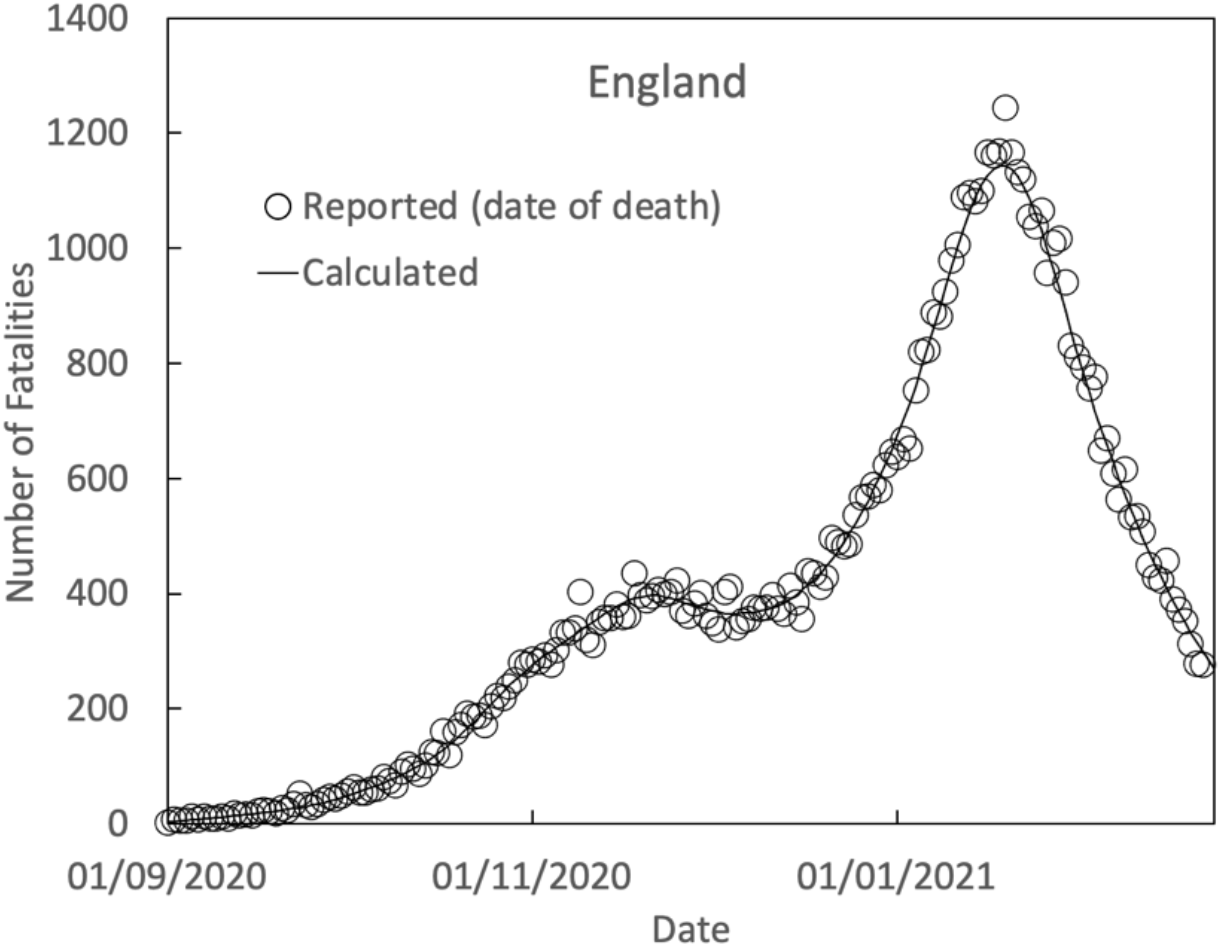
Comparison between the sum of all calculated waves of the regions compared to reported fatalities by date of death for England during the “second wave”

## 4. Discussion

The accuracy of the fitted curves to reported fatality rates, with R^2^ values approaching 100% against the ideal, strongly supports the validity of the current model and it reveals detail that would otherwise remain hidden or considered purely speculative. For example, in the “first wave” some regions show smoothly falling fatality rates with time, while others clearly show a multiple wave format. The strength of the model is it accounts for both types of behaviour in a unified approach.

It also accounts for the observed “exponential” rises in infection/fatality rates often quoted during the “second wave” which, when extrapolated into the future, substantially overestimated subsequent reported rates. Such apparent rises can happen when multiple waves, each increasing in magnitude with time, are added to each other and appear to be one continuous curve.

However, when the subsequent wave/waves either do not increase in size or diminish, it can lead to large errors in extrapolation. Fig.25 shows reported fatality rates as a 7-day average for England ^(1)^ plotted on an exponential y-axis. Extrapolation, using data for the first 2-3 weeks of October, leads to a prediction of close to 2,000 fatalities per day by the end of November, while the real value was around 400, and close to 70,000 by the end of January, while the real value had peaked around 1,200 by the middle of January.

**Fig. 25.**
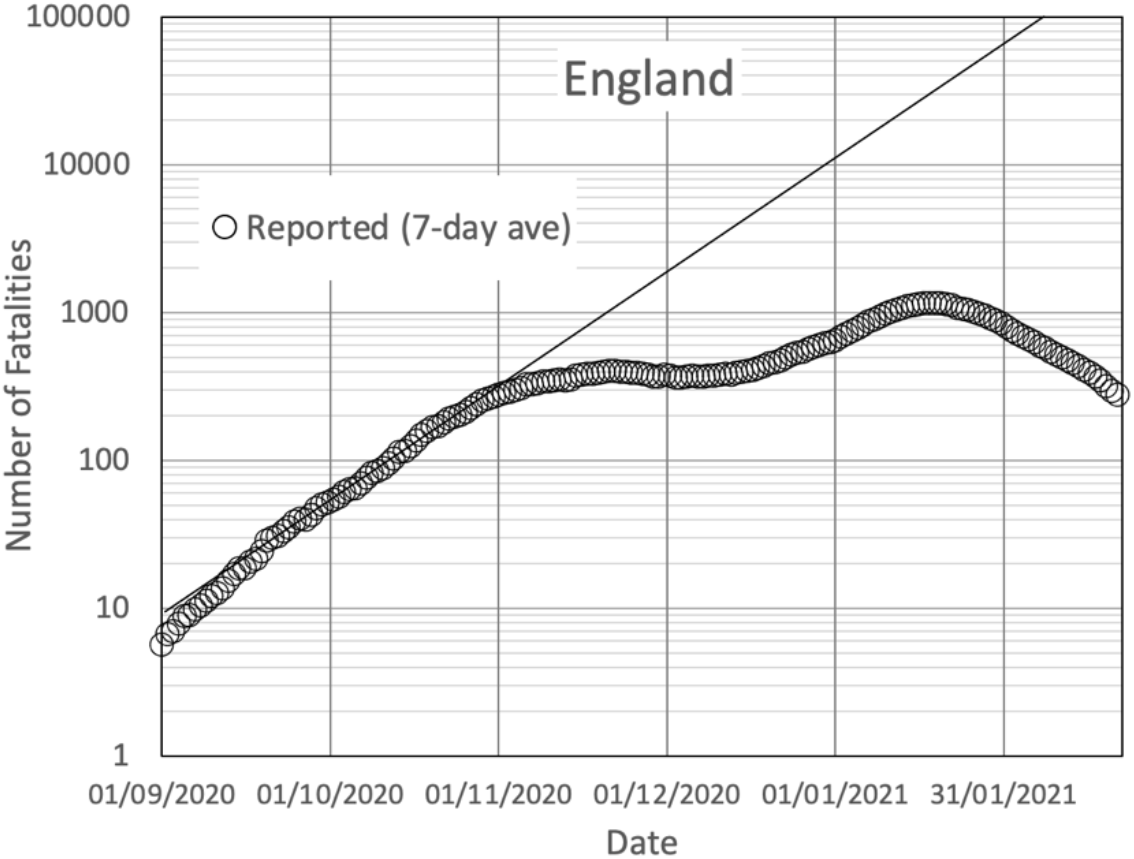
Extrapolation of the fatality curve for England in the “second wave” showing overestimation of future fatality rates by assuming exponential growth

The behaviour of the “first” and “second” waves is very different. The “first wave” consisted of a large initial spike in fatalities in all regions and was followed by a series of significantly sized secondary waves which diminished with time. London, which is the most highly interconnected of the regions, exhibited almost completely as a single wave unlike the other regions. It therefore seems reasonable to assume that the secondary waves in the other regions was due to the virus spreading into previously unaffected areas.

The “second wave” has behaved in a fundamentally different way. It began after a long period of very low fatality rates and in all regions simultaneously. Unlike the “first wave”, the “second wave” is characterised by a series of periodic waves in all regions, including London which is now exhibiting as a series of periodic waves rather than a predominantly single wave. It is also characterised by a substantial regional variation in fatality rates over time.

Fatality rates per head of population were initially highest in the Northern and Midlands regions and they first peaked during November.Rates subsequently fell until, towards of the end December, most saw quite sharp rises before peaking in January. On the other hand, in London, the South East and the East of England rates were initially quite low, but showed a continuous shallow increase until the beginning of December when they climbed sharply before also peaking in January. The South West behaved quite differently to the other regions. Its shape during the second wave was rather similar to the Midlands regions, however, the fatalities recorded per head of population remained substantially lower than all other regions.

By its very nature, the method used here is an evidence-based empirical approach and quite unlike the more theoretical approaches adopted by groups such as in Imperial College. It has the advantage in that it uses pre-existing evidence to develop an approach that can quantitatively match reported behaviour. As such, it is not within the scope of the present paper to undertake any detailed discussion on the differences between the two approaches.

However, it is noted that the success of the present model in accounting for fatality rates so accurately lies in the fact that it considers each region separately. In effect, it models England as a set of closed systems that allows regional variability of fatality rates to be taken into account. The ability to do so becomes particularly important for the “second wave”. In a more fundamental model, which includes transmission between the regions, it could be considered as a loosely coupled model with each region having its own individual characteristics yet allowing transmission between regions. But, even at this early stage, the accuracy of the current model in accounting for fatalities in England its regions does allow some specific inferences to be briefly explored.

The fact that fatality rates for London so closely matched a much-reduced ICU model during the “first wave” could be taken as indicating there was an inherent resistance to the virus in a large proportion of the population. In that light, it is further noteworthy that the similarly highly interconnected large cities New York City, Paris and Madrid also exhibit almost wholly as a single “unconstrained” wave form (Appendix 2) rather than as an ICS model wave form. In particular, daily fatalities in Madrid had fallen to almost zero by the beginning of June, while in London the same occurred by July.

It might be inferred that, if there was inherent resistance to infection in sections of the population, then the R number for the population that was susceptible to the virus would be markedly higher than expected from a national average. That would appear consistent with how rapidly infections can spread through certain settings, for example, care homes, and further consistent with observed behaviour of household infections where there “…*is evidence for clustering of SARS-CoV-2 infections within households, with some households having many secondary infections while many others have none”* ^(3)^.

After a long period during the summer when fatality rates remained very low, infection rates and subsequent fatalities rose in all regions after week 35 as early autumn approached, which would appear consistent with a seasonal rise for respiratory infections.

The nature of the “second wave” is further defined by the periodic appearance of waves of fatalities in all regions and it seems no coincidence that, as the “second wave” progressed, as one wave subsided, shortly before or afterwards, a new wave began, which could be interpreted as displaying how variants are manifesting themselves. In that light, it is noted that the most predominant wave in the southern regions is the 6^th^, which can be associated with the new Kent variant ^(4)^.

## Conclusions

The current paper demonstrates that it is possible to quantitatively model the clear observation from measured infection/fatality rates that there have been many waves of infection and subsequent fatalities in the regions of England. It further provides a clear and quantitative explanation as to why exponential extrapolations which have been made during the “second wave” have invariably failed by such large margins and accounts for fatality curves which quite clearly exhibit multiple waves as well as those that appear quite continuous.

The nature of how COVID-19 has spread is briefly discussed and potential reasons are examined as to how the multiple wave form has arisen.

## Data Availability

All of the data referenced are in the public domain

https://coronavirus.data.gov.uk/

https://web.stanford.edu/~chadj/Covid/Dashboard.html

## About the author

The author is a researcher and scientific modeller in the physical sciences and has written this paper as a contribution to the further understanding of the spread of COVID-19.

## Appendix 1. Fitted curve for London Fatalities per day based on a 7-day average of reported data ^(1)^

The curve below is used as a “master wave”. It is based on the reported 7-day average for London and, fitted to the reported data using a combination of fitting to a polynomial function and manual adjustment to provide a smoothly changing curve. While the fitted curve matches the reported fatalities for London very well, it would clearly be preferable for a more meaningful wave function to be developed to match the behaviour of London.

|  |  |
| --- | --- |
| 09/03/2020 | 0.00 |
| 10/03/2020 | 1.10 |
| 11/03/2020 | 2.46 |
| 12/03/2020 | 4.20 |
| 13/03/2020 | 6.41 |
| 14/03/2020 | 9.16 |
| 15/03/2020 | 12.55 |
| 16/03/2020 | 16.64 |
| 17/03/2020 | 21.52 |
| 18/03/2020 | 27.26 |
| 19/03/2020 | 33.95 |
| 20/03/2020 | 41.66 |
| 21/03/2020 | 50.47 |
| 22/03/2020 | 60.46 |
| 23/03/2020 | 71.71 |
| 24/03/2020 | 84.31 |
| 25/03/2020 | 98.32 |
| 26/03/2020 | 111.25 |
| 27/03/2020 | 123.76 |
| 28/03/2020 | 136.62 |
| 29/03/2020 | 149.36 |
| 30/03/2020 | 161.51 |
| 31/03/2020 | 172.61 |
| 01/04/2020 | 182.17 |
| 02/04/2020 | 191.00 |
| 03/04/2020 | 198.00 |
| 04/04/2020 | 204.00 |
| 05/04/2020 | 206.50 |
| 06/04/2020 | 207.60 |
| 07/04/2020 | 207.00 |
| 08/04/2020 | 204.50 |
| 09/04/2020 | 199.00 |
| 10/04/2020 | 191.50 |
| 11/04/2020 | 184.31 |
| 12/04/2020 | 174.21 |
| 13/04/2020 | 164.38 |
| 14/04/2020 | 154.83 |
| 15/04/2020 | 145.55 |
| 16/04/2020 | 136.55 |
| 17/04/2020 | 127.83 |
| 18/04/2020 | 119.38 |
| 19/04/2020 | 111.22 |
| 20/04/2020 | 103.33 |
| 21/04/2020 | 95.73 |
| 22/04/2020 | 88.41 |
| 23/04/2020 | 81.38 |
| 24/04/2020 | 74.63 |
| 25/04/2020 | 68.17 |
| 26/04/2020 | 61.99 |
| 27/04/2020 | 56.11 |
| 28/04/2020 | 50.51 |
| 29/04/2020 | 45.21 |
| 30/04/2020 | 40.20 |
| 01/05/2020 | 35.49 |
| 02/05/2020 | 31.07 |
| 03/05/2020 | 26.95 |
| 04/05/2020 | 23.12 |
| 05/05/2020 | 19.60 |
| 06/05/2020 | 16.37 |
| 07/05/2020 | 13.45 |
| 08/05/2020 | 10.83 |
| 09/05/2020 | 8.51 |
| 10/05/2020 | 6.50 |
| 11/05/2020 | 4.79 |
| 12/05/2020 | 3.39 |
| 13/05/2020 | 2.30 |
| 14/05/2020 | 1.52 |
| 15/05/2020 | 1.05 |
| 16/05/2020 | 0.60 |
| 17/05/2020 | 0.22 |
| 18/05/2020 | 0.00 |

## Appendix 2. Reported fatality rates for London, New York, Paris and Madrid

Figures 26(a-d) show reported fatalities using a 7-day average during the “first wave” for the four major cities, London, New York, Paris and Madrid fitted with a single wave as calculated by the present model. Other than London, reported fatalities are taken from the study of Fernández-Villaverde and Jones ^(5)^ and downloaded from the web page shown in ^(6)^.

**Fig 26.**
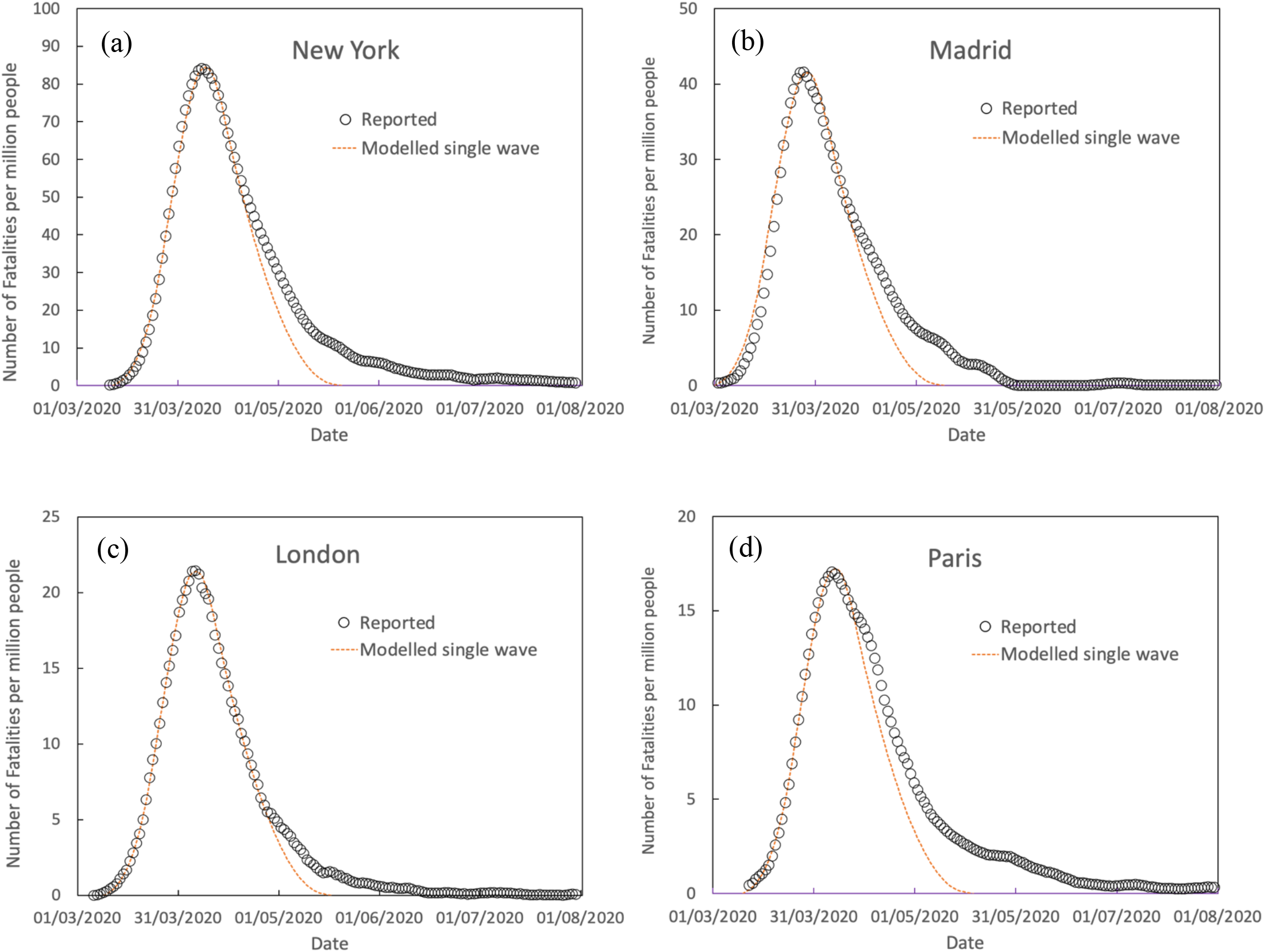
Reported daily fatalities per million people (7-day average) in comparison to a single modelled wave for (a) New York, (b) Madrid, (c) London and (d) Paris

Following the format of the source data from Fernández-Villaverde and Jones, plots are shown as fatalities per million of population, with the results for London converted from ref.1 using the population for London as provided by the UK’s Office for National Statistics. What is clear from the plots is that, while all cities exhibit predominantly as a single wave, there is a significant variation in fatalities per head of population between them. New York had, by far, the most severe death toll in comparison to the European cities, while Madrid fared worse of the European cities. Of further interest is that the daily fatality rates for London and Madrid had fallen to almost zero by June/July.

## Footnotes

1. UK Government website for data and insights on Coronavirus (COVID-19) https://coronavirus.data.gov.uk/

2. N.M. Ferguson et al., Report 9: Impact of non-pharmaceutical interventions (NPIs) to reduce COVID-19 mortality and healthcare demand, (Imperial College COVID-19 Response Team, 16 March 2020)

3. Z.J. Madewell *et al*., “Household Transmission of SARS-CoV-2A Systematic Review and Meta-analysis”, JAMA Network Open. 2020;3(12)

4. T. Kirby, “New variant of SARS-CoV-2 in UK causes surge of COVID-19”, The Lancet, vol.9(2), (2021), E20-E21

5. J. Fernández-Villaverde and C.I. Jones, “Estimating and Simulating a SIRD Model of COVID-19 for Many Countries, States and Cities”, https://web.stanford.edu/~chadj/sird-paper.pdf

6. https://web.stanford.edu/~chadj/Covid/Dashboard.html

